# Investigating the Relation between Functional Road Class and Hospital Diagnosed Injury Severity for Crashes in Maryland

**DOI:** 10.1101/2025.09.23.25335901

**Authors:** Adewumi Adegboye, Kartik Kaushik

**Affiliations:** Department of Epidemiology and Public Health, University of Maryland School of Medicine, Baltimore, Maryland, 21201; National Study Center for Trauma & EMS, Shock Trauma and Anesthesiology Research, University of Maryland School of Medicine, Baltimore, Maryland, 21201

**Keywords:** Motor Vehicle Crash, Injury Severity, Blood Alcohol Content, Functional Road Classification, Data Linkage, Injury Severity Score, Abbreviated Injury Scale

## Abstract

The true toll of Motor Vehicle Crashes (MVC) lies in mortality and injuries that force lifelong changes, impacting mental health, occupation, means of attaining livelihood for self and family, and long-term chronic suffering. However, limited emphasis has been placed on morbidity as compared to mortality from MVC. This paper presents results from modeling the association between injury severities, Blood Alcohol Content (BAC), and roadway classes using data from 2016 – 2021 obtained from the R Adams Cowley Shock Trauma Center (STC) registry linked with police crash reports and the Functional Road Class (FRC) data at the patient level for Maryland. The record-level linkages between the STC and police data were produced by a linear combination of edit distance scores between common variables in either dataset, while geospatial proximity linkage was used to combine FRC data with incident locations. Ordered logistic regression model with partial proportional odds was used for determining impact of exogenous variables on injury severities. The model incorporates BAC obtained from blood screens at the time of arrival to the hospital. Moreover, the analytics presented use globally accepted consensus derived injury severities based on hospital diagnoses. The three elements taken together are unique to this study and are not replicated in existing literature, although they have been studied individually. Results show that facilities from major arterials to local roads have a higher risk for more severe injuries and longer hospital stay compared to facilities like interstates and freeways. Moreover, the risk of injuries that can cause permanent lifestyle changes due to disabilities is also higher on those roads.

## 1. INTRODUCTION

While there has been, and continues to be, significant emphasis placed on mortality resulting from Motor Vehicle Crashes (MVC), injuries, no matter how severe, are often overlooked. The complexity of the issue lies in the multifaceted nature of injuries. Classifying and assessing the short- and long-term impacts and costs of injuries can vary by injury types, severities, and individuals, even for similar injuries caused by similar mechanisms. Pain tolerances, required interventions, prognosis, and post-discharge recovery all vary on a multitude of factors, from access to advanced medical and support services to personal health, complications, and comorbidities. Moreover, a joint investigation of crashes and injuries is inherently multidisciplinary, requiring subject matter expertise from engineering, law enforcement, and medicine. Such interdisciplinary teams are rarely formed, reducing the likelihood of conducting research similar to the study presented in this paper.

This paper describes results from a study to investigate whether an association exists between roadway types, blood alcohol content, patient demographics, and injury severity. It is hypothesized that with higher emphasis and funding for active transportation, increased environmental consciousness, and availability of electrified personal micromobility vehicles, mixed-use roadways are more dangerous than large-capacity interstate roads. The results of this study adds to the mounting evidence confirming the hypothesis, urging research, treatments, and funding to improve the infrastructure of mixed-use facilities to accelerate Vision Zero to eliminate all fatalities and serious injuries. The study combines individual patient data mined from medical charts with crash records from the Maryland State Police (MSP) and the Functional Road Class (FRC) data obtained from the Highway Performance Monitoring System (HPMS) dataset produced by the Maryland Department of Transportation (MDOT). The data are linked at the individual patient and crash level, and the injury severities are then analyzed using Partial Proportional Odds Model (PPOM), a type of Ordered Logistic Regression (ORL) where disproportionate exogenous variables are allowed separate coefficients for each ordered level of the endogenous variable.

### 1.1. Literature Review

Many published works discuss the use of proportional and partial proportional odds models in the context of injury severities (*1 – 9*). The various published works using partial proportional odds models generally have shown good performance of the models in establishing a relation between the explanatory variables and injury severities, albeit the works investigate specific types and instances of crashes or specific types of roadway users (*1 – 9*). The authors of this study have been unable to find any published literature that simultaneously uses global consensus derived injury severities based on medical imaging and diagnosis modeled as a function of the roadway class and blood alcohol content obtained from blood tests at admission along with basic demographic variables.

Traditional analytics into crash severity research was focused primarily on volumes and speeds and urban and rural roadways, showing how moderate volumes and higher speeds contribute to more severe crashes (*10 – 13*). Research interest was also bestowed on the demographics, social backgrounds, and choices made by drivers regarding the risks of injury severities, often finding young males and habitual speeders to be at the highest risk for severe injuries (*14 – 16*). Stiles et al. (2021) comprehensively uses road classification as a predictor of injury severity; however, they use severities recorded by the police (*17*). Although very limited, earlier studies usually defined road types and characteristics to suit the research question at hand, often restricting the study scope to a single roadway type or even a corridor and lack generalizable results (*18*).

The robust roadway design process, used by the Federal Highway Administration (FHWA) and all state Departments of Transportation (DOT) in the United States, accounts for roadway type implicitly by tying it rigidly to the designed volumes and speeds. Safety is affected by controlling the speeds given expected volumes and use of the roadways (*19*). However, given the modern rapid adoption of active transportation, the large array of personal micromobility vehicles that emphasize speed, convenience, and effortless travel, and the quick build projects many cities and localities undertake, the established paradigm is quickly shifting (5 – 9, *20, 21*). This is evidenced by the decade-long rise in pedestrian fatalities that are now exacerbated by micro-mobility vehicles (*5 – 9, 20, 21*). A similar divide can also be seen across urban and rural regions, where the rising trend of fatalities in urban spaces far outpaces the trends in rural areas (*21*).

A 2022 study by the National Highway Traffic Safety Administration (NHTSA) found that 56% of all serious injury and fatal crashes involved alcohol or drugs, with 20% of cases involving multiple impairing substances. Further, the study showed that cannabis was detected in 25% of cases followed by alcohol in 23% of cases (*22*). While in 2006, 28% of all driver fatalities were impaired by drugs, the share ballooned to 44% by 2016. Moreover, polydrug use among driver is also growing, increasing the risks of impaired driving (*23*). By 2020, 30% of all traffic fatalities in the US involved alcohol, resulting in an alcohol involved fatality every 45 minutes (*24*). 21.9% of all drivers in 2021 used illicit drugs, primarily marijuana with 52.5 million users (*25*).

All studies referenced so far rely on the injury severities recorded by the police, which are seldom medically accurate as the police are neither medically trained nor have access to advanced diagnostic equipment (*26, 27*). The use of medical diagnoses-derived injury severity scales has mainly been featured in papers that compare them to the injury severities captured by law enforcement officers (*26, 27*) and explore medical nuances of the kinds of injuries that result from transportation crashes. This study aims to investigate injury severities primarily as a function of the roadway class and impairment by alcohol in addition to basic demographics.

## 2. DATA DESCRIPTION

### 2.1. STC Trauma Registry Data

Patients admitted to the R Adams Cowley Shock Trauma Center (STC) at the University of Maryland Medical Center (UMMC) in Baltimore form the base population of this study. The STC is among the United States’ busiest trauma centers and is the only designated Primary Adult Resuscitation Center (PARC). PARC designation indicates round-the-clock access to advanced surgeons, anesthesiologists, medical personnel, and facilities that go beyond the requirements of a Level 1 Trauma Center. The STC admits and treats about 2,000 patients from MVCs each year, especially the most severely injured. Patients can arrive at the STC by land or air ambulance only, based on a statewide triage protocol. Injuries to each body region recorded in the patient’s medical chart were scored according to the Abbreviated Injury Scale (AIS) manual and then combined for a total Injury Severity Score (ISS) by a dedicated team at the hospital.

The AIS is a globally accepted consensus derived injury severity score, while the ISS is a combination of AIS from multiple body regions to score poly trauma. AIS subdivides the human body into eight regions – head, face, neck, chest, abdomen, spine, upper extremities (hands), and lower extremities (legs) – with six levels of possible severities for each. The possible severities of injuries to each body region range from 1 (mild superficial injuries) to 6 (maximal injury). Injuries to certain body regions, such as the extremities, cannot attain a severity of 6 as they are seldom life-threatening. The ISS is obtained by adding the squares of the AIS score for three worst injured body regions and ranges from 1 to 75, where a score of 1 – 8 is considered as mild injury, 9 – 15 as moderate injury, 16 – 24 as serious injury, 25 – 74 as severe injury, and 75 as maximal. An AIS of 6 for any body region automatically scores 75 on the ISS (*28 – 32*).

The STC also conducts a comprehensive toxicology screening on every patient at admission. The primary reason for the toxicology screening is to ensure that medications given to the patient do not conflict with or exacerbate the effects of any existing drugs in the system. A total of 14 drugs are screened along with alcohol. Since impairment is not directly assessed at the time of drug screen, it is not immediately possible to determine if the screen results indicate impairment or residual metabolites. Therefore, this study includes only the values of the BAC results, although future studies will include results of other drugs and expert opinion on the likelihood of impairment caused by the drug screen results.

The STC maintains a data registry with details about patients treated at the STC. Details include patient demographics, admission and discharge times and dispositions, arrival details like place of origin, mechanism of injury, etc., diagnostic data, hospital course information like procedures performed, diagnostic tests conducted, lab results, vitals, and so on. The population of 10,883 patients used in this study was obtained from the STC registry, defined as direct admissions from the scene with an external cause of injury due to a land transportation crash. All patients were admitted from crashes occurring in Maryland between January 1, 2016, and December 31, 2021.

The registry was queried for patient demographics on all patients in the population. Additionally, the computed ISS and the BAC results were obtained. The BAC screen lists the exact tested amount of alcohol in blood as a percent value. The results were categorized into negative, below the per se Driving Under Influence (DUI) limit of 0.08, and above the per se DUI limit. Categorizing the variable allows direct understanding of the differences in injury severities relative to existing laws.

### 2.2. MSP Automated Crash Recording System (ACRS) Data

The MSP use the Automated Crash Recording System (ACRS) to record all traffic crashes in the state of Maryland, which is one of the few states in the US with a standardized, electronic, statewide traffic crash data collection system. All unintentional traffic crashes with police response in the state of Maryland are recorded in ACRS, together with the geocoordinates of the crash. Crash records of patients in the study population were searched in the ACRS database. The circumstances of the crash, including the geocoordinates of the location of the crash, are recorded for every matching patient.

Levenshtein textual distance (*33*) was used to link the datasets given recording differences, missing data, and errors over an array of common columns. A linear combination of the distance scores (higher the better) across the common columns were used to identify the best candidate match record. Over all the patients in the dataset, match scores below 65^th^ percentile were rejected as non-matches. Scores above 85^th^ percentile were selected as matches. Records with scores in between were manually reviewed. Since both datasets are mostly recorded by hand, exact match criteria are difficult to satisfy. The common columns included data classified as Personal Identifiable Information (PII), including names, dates of birth, and addresses. The search for a matching police record for each patient was limited to police recorded crash times 4 hours before and 2 hours after patient admission.

Patients without a corresponding record in the police crash report were excluded from subsequent analysis. The number of patients remaining in the study population with police crash records is 8,912, about 18% less than patients identified in the STC registry. A zip code level heatmap of the place of injury for patients with police crash records is shown in **figure 1**. A log (base 10) scale is used to color the zip codes in F**igure 1** to account for large differences in the absolute numbers of patients admitted from each zip code.

**FIGURE 1:**
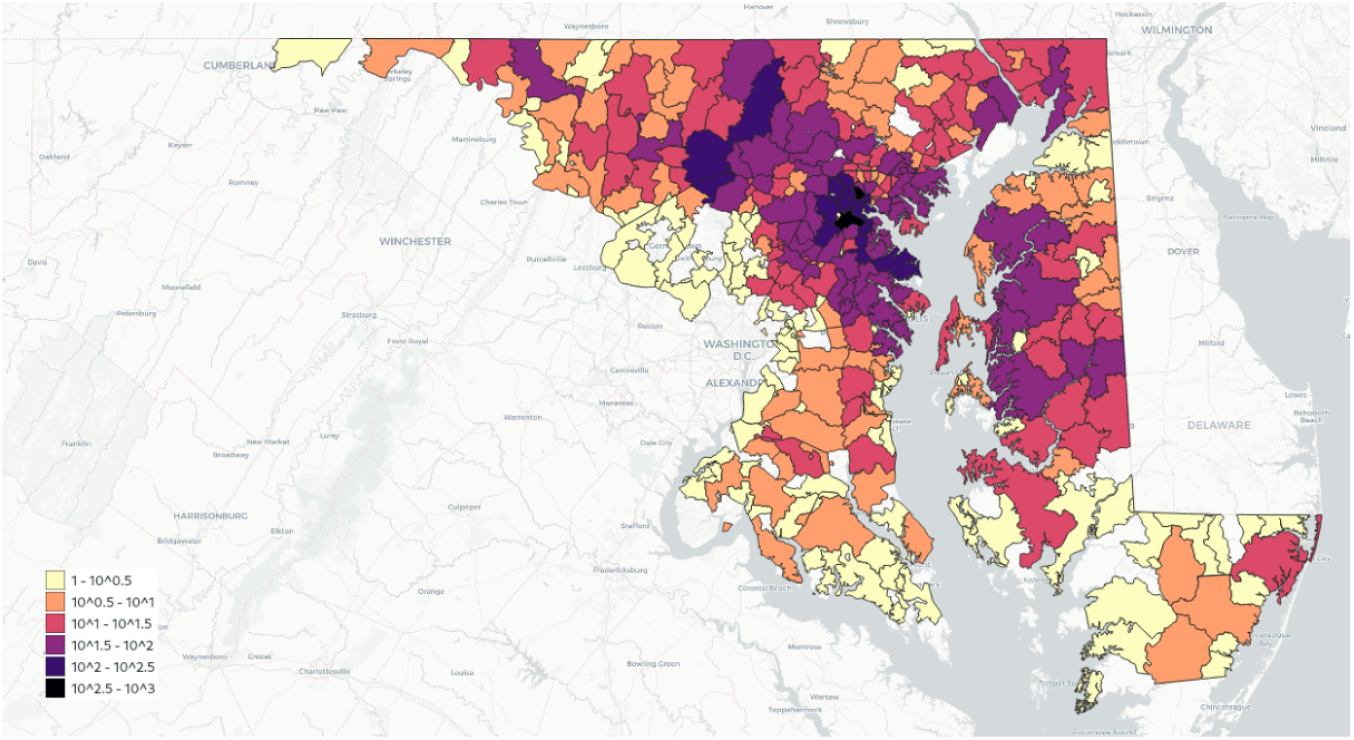
Zip code level heat map of patients with ACRS crash records, N = 8,912.

### 2.3. MDOT FRC Roadway Layer

The FHWA, through the HPMS, classifies roadways into seven different classes based on the type of service provided. The FRC is also an ad-hoc measure of the friction expected on roadways, with classes 1 and 2 representing Interstates and Freeways, being synonymous with faster, high-capacity facilities designed for speedy movement of people and goods with minimal friction to traffic flow. Classes 5 through 7 provide access to secondary roadways and destinations such as businesses, services, and local residences. The intermediate classes – 3 and 4 – function as primary and secondary arterials, connecting the interstates with the local roads. While these classes can also provide access to destinations they are more optimized for faster traffic flow. Each state classifies the roadways in its jurisdiction in accordance with the HPMS classes.

The HPMS roadway layer for Maryland was obtained from the MDOT Open Data Portal (https://data-maryland.opendata.arcgis.com/pages/mdot). The data contains the road geometries in a geospatial shapefile, together with the road numbers, road names, and FRC of the roadway. A spatial join was performed to attribute the crash location obtained from the ACRS data to a roadway in the HPMS layer. Roadway segments that intersected a buffer of 150m (492ft) around the crash geocoordinate were selected as candidate segments for the spatial linkage. Additional weight was applied to the Levenshtein textual distance between the road names and numbers recorded by the police and of the roadway in the HPMS data (*34, 35*). The closest segment in terms of spatial and textual distance was selected as the candidate segment. Due to geolocation being considered PII, a pictorial representation of the linkage cannot be provided. Only two records failed to conflate with the correct roadway segment and so were removed from the final dataset used for the analysis. These crashes happened inside tunnels and so were poorly geolocated.

### 2.4. Final Dataset

The linked dataset contains a record for each patient, with 8,910 patients in total. Records with missing values in the selected variables were excluded, yielding 8,203 observations in the final dataset. The selected variables are:

- Patient demographics: age, gender, and admission time stamp
- Patient injury severities: Combined ISS scores, classified as 1 – 8 mild, 9 – 15 moderate, 16 – 24 serious, 25 – 74 severe, and 75 maximal.
- Patient Blood Alcohol Content: classified as negative, below per se (0.08), and above per se
- Crash circumstances: Person type as driver, passenger, or non-motorist, crash location, and crash time stamp.
- Functional Road Classification: 1 – Interstate, 2 – Freeway, 3 – Principal Arterial, 4 – Minor Arterial, 5 – Major Collector, 6 – Minor Collector, 7 – Local Road.

## 3. METHODOLOGY

Cumulative or ordered logistical regression (logit) models are frequently used to make inferences about ordinal response data (*36*). Injury severities orderly increase from minor to maximal injuries. Therefore, cumulative logit models are perfectly suited to model injury severities. Ordinary ordered logit models assume that the proportions of effects between adjacent ordered levels are maintained. This assumption is called the proportional odds assumption or the parallel lines assumption. However, injury severities do not scale in proportion to the external causes and contributing factors. Consider, for example, the risk of fatality as a function of vehicle speed. As speed increases, the risk of fatality increases more than proportionally (*37*). Consequently, the parallel lines assumption needs to be relaxed. The opposite of the proportional odds model is the non-proportional odds model, which is equivalent to a generalized ordered logit model. An intermediate model is the partial proportional odds model where the assumption of proportionality is only relaxed for those exogenous variables that scale non-proportionally to the response variable (*38*). All three models were examined for this paper on the same set of data. All statistical tests were performed by using SAS software release 9.4 (SAS Institute Inc, Cary, NC).

### 3.1. Proportional odds model (parallel lines assumption)

The proportional odds model (POM) described by McCullagh (*36*) is the most accepted model for ordinal logistic regression (*39*). The POM can be referred to as cumulative or ordinal logit model. The feature of the POM is that the odds ratio for a predictor can be interpreted as a summary of the odds ratios obtained from separate logistic regressions using all possible cut points of the ordinal outcome (*40*). Further, the probabilities of a higher cut point are cumulative over the lower cut points. The underlying assumption of proportionality gives that all jumps are equidistant from each other by constraining the model coefficients to be equal across all cut points. The equation for cumulative logit model is (*40*):

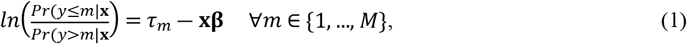

where *m* is the ordered response category, *τ* is a cut point, **x** is a vector of exogenous variables, and **β** is a vector of coefficients.

The POM was fitted to the data used in this paper. However, the injury severity scale is non-linear as an increase in severity more than proportionately reduces the likelihood of survival (1 – 5). Consequently, a simple cumulative logit with proportional odds was found to be not applicable to the data as the proportional odds null hypothesis was statistically rejected (*p* < 0.001). It has further been shown that a stringent significance threshold, of up to 0.01, might be better suited to this class of models due to the number of tests being conducted (*29*). A nonsignificant test is taken as evidence that the logit surfaces are parallel and that the odds ratios can be interpreted as constant across all possible cut points of the outcome. Alternative models include a partial proportional odds model, a non-proportional odds model, or a dichotomous ordinal outcome at the cost of statistical power (*41*).

### 3.2. Partial Proportional Odds Model

The partial proportional odds model (PPOM) is an intermediate method that connects the gap between ordered logit (POM) and generalized ordered logit models (*38*). These models allow for some independent explanatory variables to affect each level of the response variable differently, while other independent variables adhere to the proportional odds assumption. Mathematically, these models can be represented as (*40*):

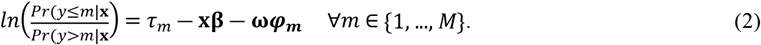

The difference between **Equation 1** and **Equation 2** is the separation of the exogenous variables into two vectors **x** and **ω**, with coefficients **β** that are fixed and **φ** that vary across cut points, respectively. The rest of the terms are the same as above. This model allows for coefficients of explanatory variables with relaxed proportionality assumptions to vary across all cut points, while retaining the proportionality assumption where appropriate.

Interpreting the results of intermediate categories need caution as the signs of the coefficients may not be consistent to determine the direction of the effect (*38 – 41*). Consequently, the marginal effects of the variables were used in this study for interpreting the results of the model. The marginal effects estimated for the independent variables measure how changes in the independent variables affect injury severity at different levels. The variables that do not require relaxation of parallel lines assumption are interpreted similar to coefficients of any ordinal model.

### 3.3. Non-Proportional Odds Model

The Non-Proportional Odds Model (NPOM) is essentially a generalized ordered logit model, where the efficiency is sacrificed to a less parsimonious model. NPOM fits coefficients to every independent variable at each cut points, thus requiring estimation of more parameters. In the presented study, the outcome variable has five levels (four cut points) and six independent variables requiring 28 parameters. On the other hand, a POM would only require 10 parameters. The larger number of parameters causes the NPOM to be less efficient and more prone to over fitting. However, in order to facilitate comparisons, NPOM was fitted to the data and is presented in this paper. An NPOM can be given as (*40*):

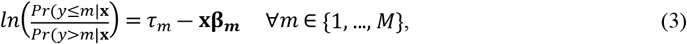

where the notations mean the same as **Equations 1** and **2** above.

## 4. RESULTS

The final dataset contained 8,203 observations. The unit of observation is an admission at the STC. The bivariate distribution of all explanatory variables used in the modeling are presented in **Table 1**. The total frequency of patients in different parameter categories and the proportions of different injury severity levels for each variable category are provided in the table. Note that row percentage of different injury severity levels for each variable category are presented.

**TABLE 1:**
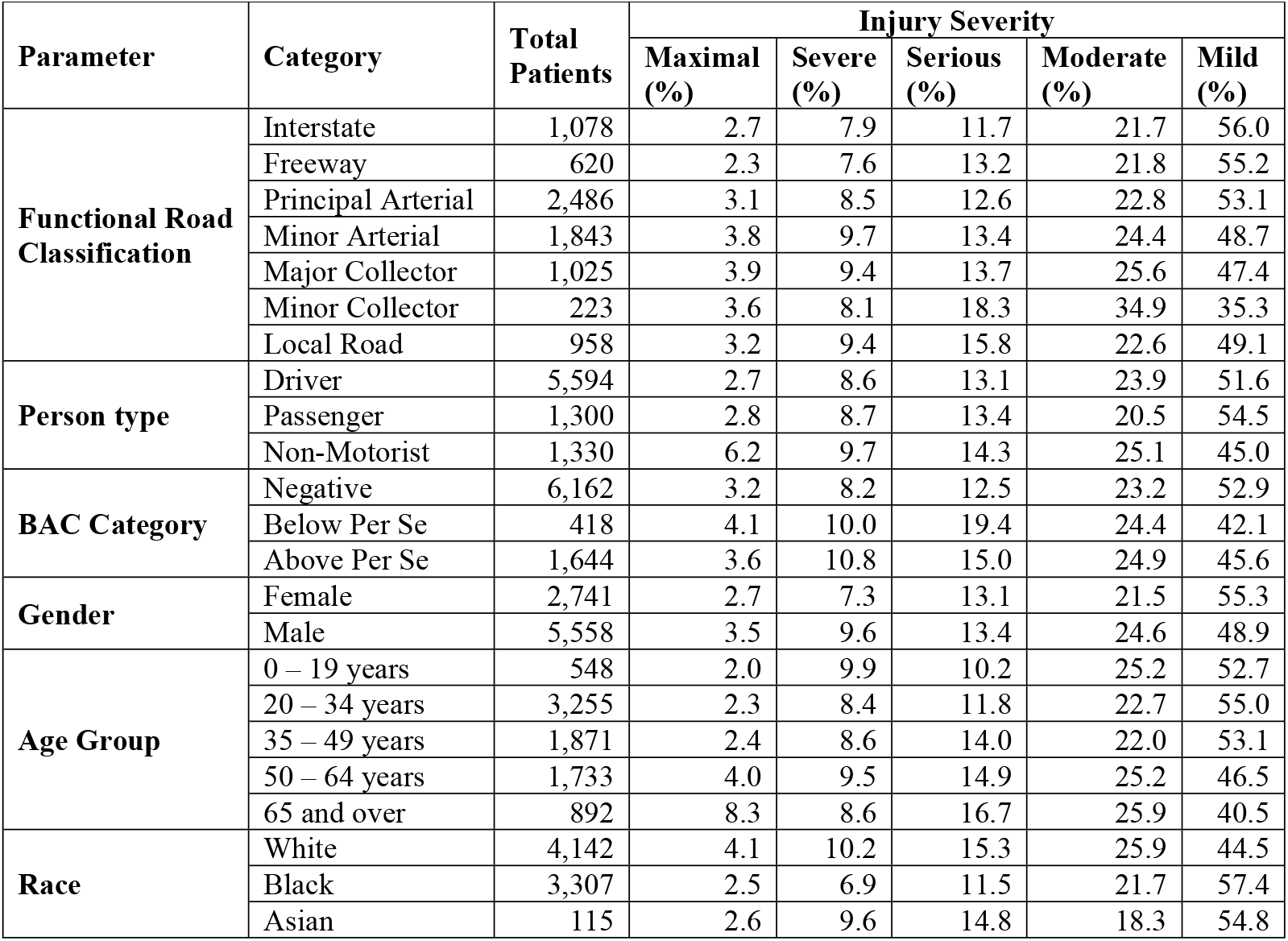
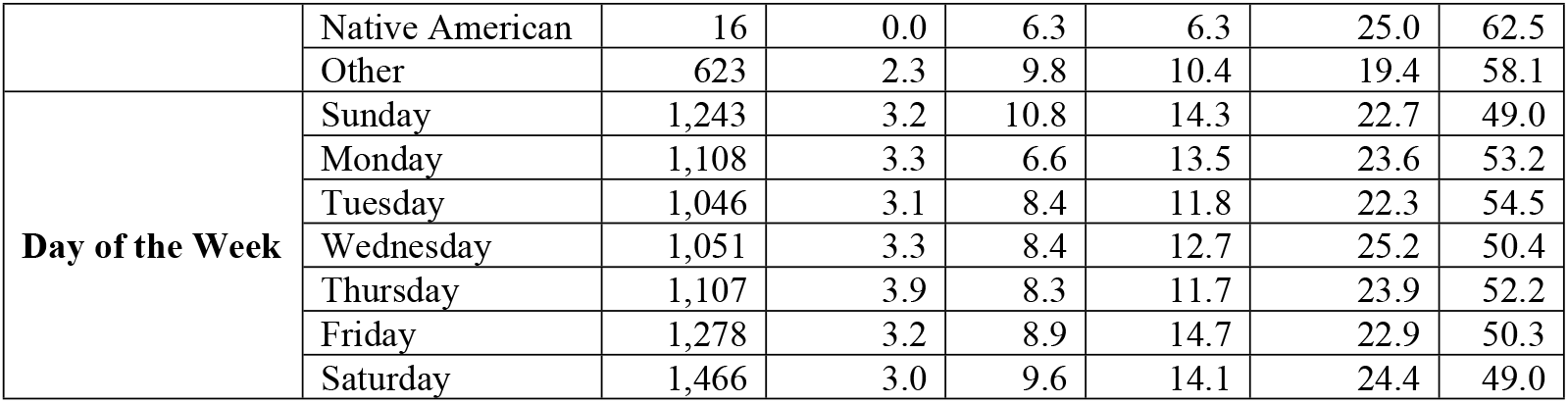
Bivariate relations of variables by injury severity level.

The proportions are the unadjusted odds and at the outset provide the interactions between different variables and the outcome of injury severity. For example, it can be seen that the majority of crashes occur on Principal Arterials, and the least on Minor Collectors. However, the odds of suffering a mild injury is more than not for Interstates, Freeways, and Principal Arterials, indicating that the other roadway classes contribute often to worse injuries than these large volume and higher speed facilities. Similarly, the proportion of mild injuries drops as a person ages, or is impaired, or is a non-motorist, or is male, and so on. The model presented later in the paper adjusts these odds so that one may interpret the contribution of a parameter to the outcome.

**Figure 2** shows the distribution of age and the share of gender given FRC. The figure shows minimal variation across the functional classes for either of the two variables. This result is expected as a difference in usage of a road class by a group of people is unexpected.

**FIGURE 2:**
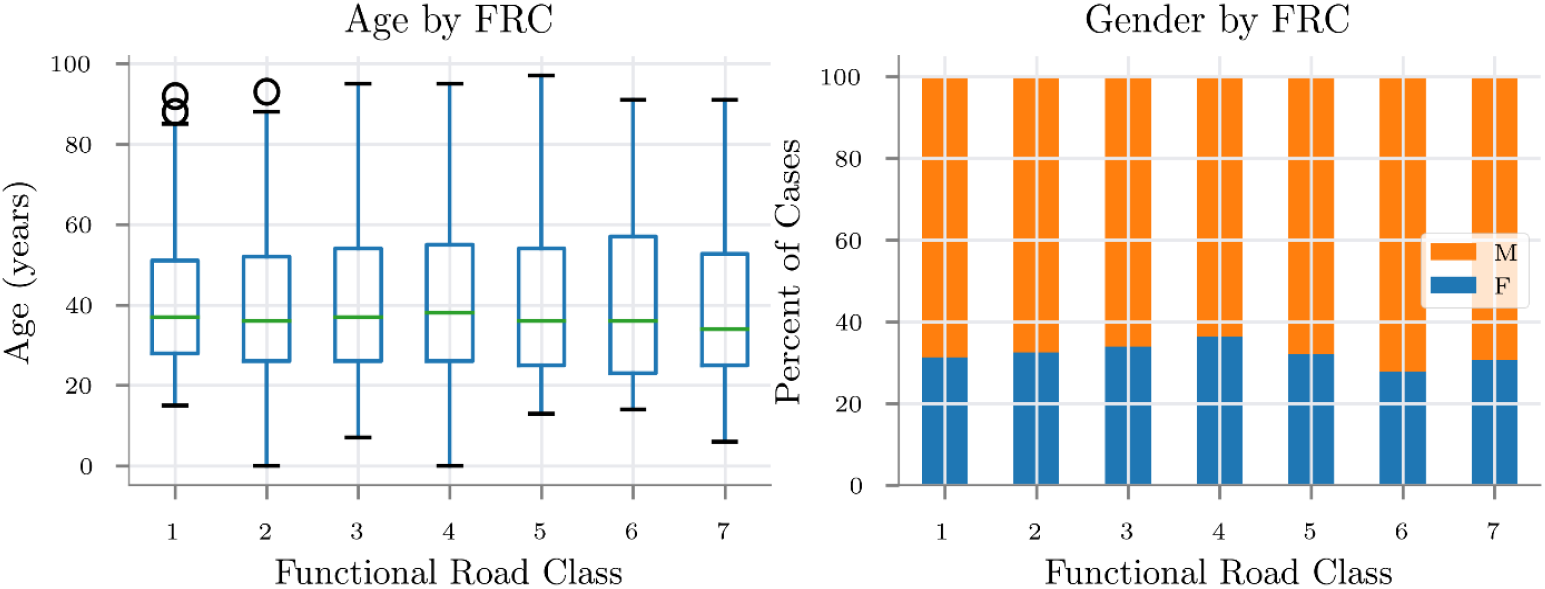
Patient demographics by FRC. 3a: Age and 3b: Gender.

The proportional odds assumption is assessed for each variable used in the model. Additionally, plots of the empirical cumulative logit function are shown as a visual guide of the parallel lines assumption in **Figure 3**. For the assumption to not be violated, the curves of the independent variable plotted against the empirical logits should be parallel. The lines in the figure are empirically estimated at the cut points, and consequently are one less than the number of levels of injury severity. In the plot these levels are denoted as 1, 2, 3, and 4 (see legend of the plots).

**FIGURE 3:**
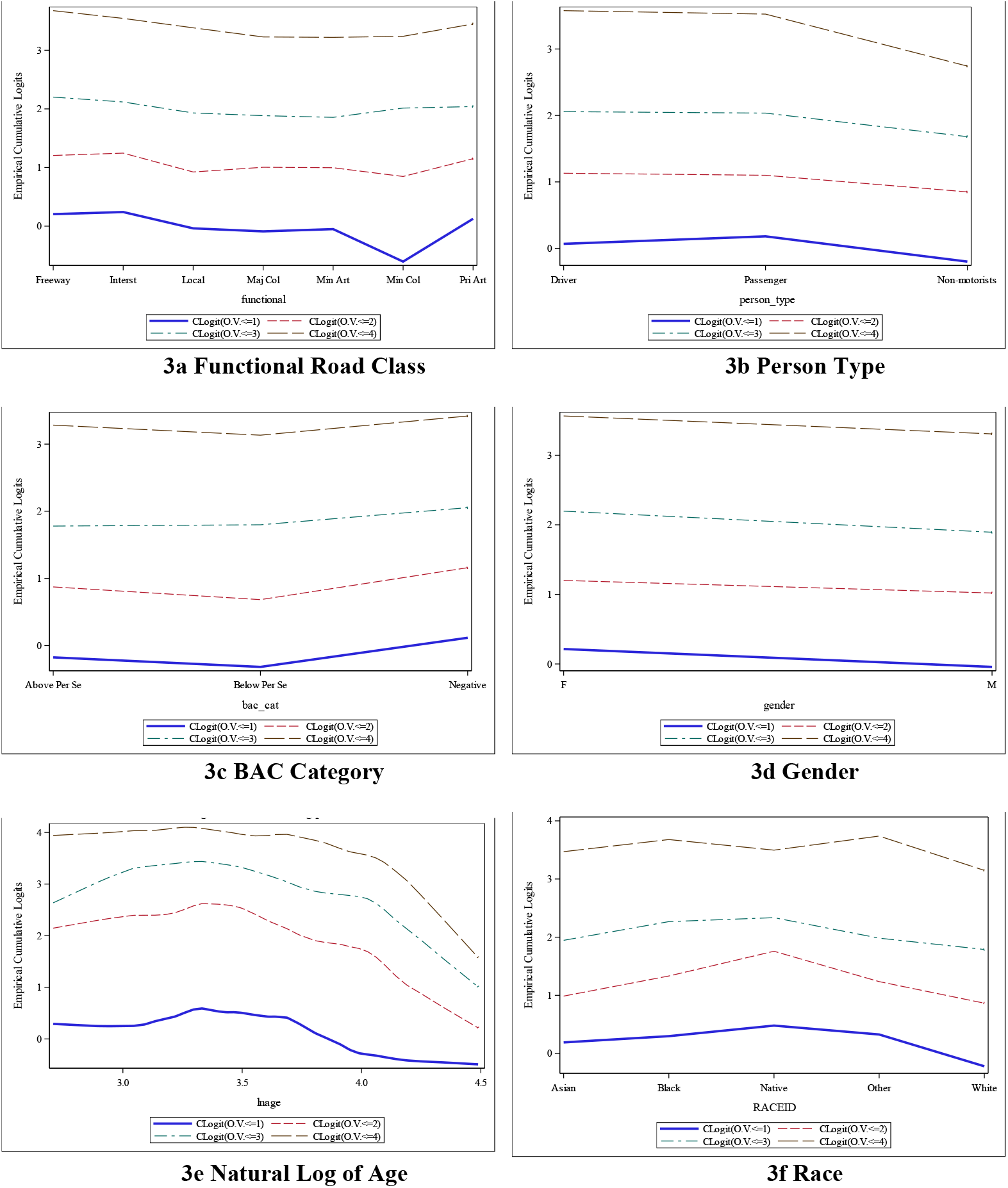
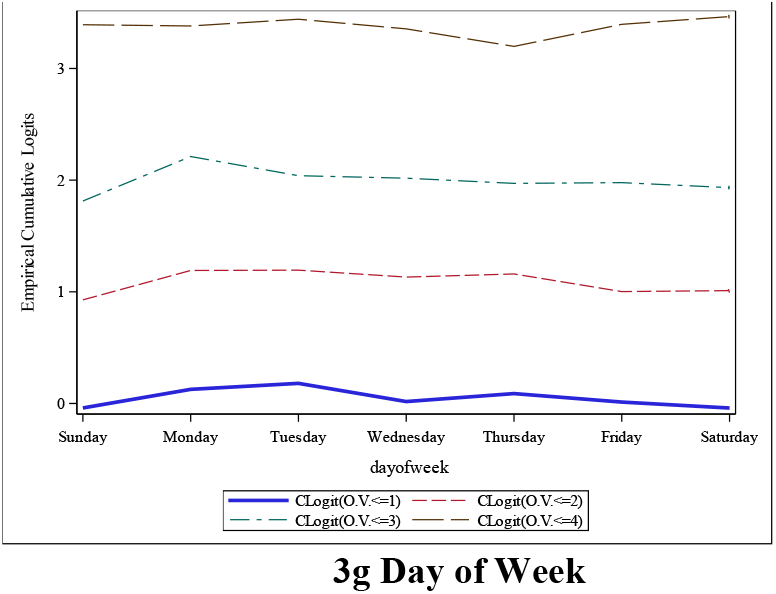
Graphical Test for Proportional Odds using Empirical Cumulative Logits.

In **figure 3a**, the cumulative logits for FRC are plotted. The test for proportional odds assumption was not violated (*p* = 0.393). On the other hand, **Figure 3b** shows the logits for person type and indicates a significant deviation from the assumption (*p* < 0.001). Therefore, person type is modeled with the parallel lines assumption relaxed. Similarly, the parallel lines assumption was not violated for BAC category (**figure 3c**, *p* = 0.582), gender (**figure 3d**, *p* = 0.085), race (**figure 3f**, *p* = 0.270), and day of week (**figure 3g**, *p* = 0.502) variables, while it is violated for the natural log of age (**figure 3e**, *p* < 0.001) variable. Consequently, FRC, BAC category, gender, race, and day of the week are modeled with a constant coefficient across all cut points, while the coefficients of person type and age are allowed to vary. Coefficients of model intercept always vary at the cut points.

**Table 2** shows the estimates produced by the partial proportional odds model. The table includes the coefficient estimates, the standard error in the estimate, the odds ratios compared to the reference, and the 95% Wald confidence intervals for the odds ratios. Each categorical variable presented in **Table 2** has a reference value as indicated in the table. Moreover, the probabilities are cumulative over the lower levels and the population group of Mild injuries acts as the reference for the model. Coefficient estimates are marked with one, two, and three asterisks to indicate p-value significance, obtained from the Wald Chi-Squared test at 0.05, 0.01, and 0.001 levels, respectively.

**TABLE 2:** Maximum Likelihood Estimates of the Partial Proportional Odds Model.

| Parameter | Category | ISS Group | Estimate | Standard Error | Odds Ratio Estimate | 95% Wald Conf Limits |  |
| --- | --- | --- | --- | --- | --- | --- | --- |
| Intercept |  | Maximal | − 7.29 *** | 0.59 | — | — | — |
|  |  | Severe | − 2.97 *** | 0.31 | — | — | — |
|  |  | Serious | − 2.31 *** | 0.24 | — | — | — |
|  |  | Moderate | − 1.06 *** | 0.22 | — | — | — |
|  | Interstate |  | Reference | — | — | — | — |
| **Functional Road Class** | Freeway |  | – 0.01 | 0.10 | 0.99 | 0.82 | 1.20 |
|  | Primary Arterial |  | 0.07 | 0.07 | 1.07 | 0.93 | 1.23 |
|  | Minor Arterial |  | 0.20 ** | 0.08 | 1.22 | 1.06 | 1.42 |
|  | Major Collector |  | 0.22 ** | 0.08 | 1.25 | 1.06 | 1.47 |
|  | Minor Collector |  | 0.45 *** | 0.13 | 1.56 | 1.20 | 2.02 |
|  | Local Road |  | 0.19 * | 0.09 | 1.21 | 1.03 | 1.44 |
| **Person Type** | Drivers |  | Reference | – | – | – | – |
|  | Passengers | Maximal | 0.3 | 0.19 | 1.35 | 0.94 | 1.94 |
|  |  | Severe | 0.18 | 0.10 | 1.20 | 0.99 | 1.46 |
|  |  | Serious | 0.19 * | 0.07 | 1.21 | 1.04 | 1.40 |
|  |  | Moderate | 0.03 | 0.07 | 1.03 | 0.91 | 1.17 |
|  | Non-Motorists | Maximal | 0.86 *** | 0.14 | 2.36 | 1.79 | 3.11 |
|  |  | Severe | 0.45 *** | 0.09 | 1.57 | 1.32 | 1.87 |
|  |  | Serious | 0.35 *** | 0.07 | 1.41 | 1.23 | 1.62 |
|  |  | Moderate | 0.33 *** | 0.06 | 1.39 | 1.23 | 1.58 |
| **BAC Category** | Negative |  | Reference | – | – | – | – |
|  | Below Per Se |  | 0.45 *** | 0.09 | 1.57 | 1.31 | 1.89 |
|  | Above Per Se |  | 0.27 *** | 0.05 | 1.31 | 1.18 | 1.45 |
| **Gender** | Male |  | Reference | – | – | – | – |
|  | Female |  | – 0.24 *** | 0.05 | 0.79 | 0.72 | 0.86 |
| **Natural Log of Age** |  | Maximal | 1.03 *** | 0.15 | 2.81 | 2.09 | 3.78 |
|  |  | Severe | 0.29 *** | 0.08 | 1.34 | 1.14 | 1.56 |
|  |  | Serious | 0.37 *** | 0.06 | 1.44 | 1.28 | 1.62 |
|  |  | Moderate | 0.33 *** | 0.05 | 1.38 | 1.25 | 1.54 |
| **Race** | White |  | Reference | – | – | – | – |
|  | Black |  | – 0.49 *** | 0.05 | 0.62 | 0.56 | 0.67 |
|  | Asian |  | – 0.2 | 0.18 | 0.82 | 0.57 | 1.18 |
|  | Native American |  | – 0.76 | 0.50 | 0.47 | 0.18 | 1.25 |
|  | Other |  | – 0.44 *** | 0.09 | 0.65 | 0.55 | 0.76 |
| **Day of Week** | Sunday |  | Reference | – | – | – | – |
|  | Monday |  | – 0.19 * | 0.08 | 0.83 | 0.71 | 0.97 |
|  | Tuesday |  | – 0.23 ** | 0.08 | 0.80 | 0.68 | 0.93 |
|  | Wednesday |  | – 0.08 | 0.08 | 0.92 | 0.79 | 1.08 |
|  | Thursday |  | – 0.15 | 0.08 | 0.87 | 0.74 | 1.01 |
|  | Friday |  | – 0.09 | 0.08 | 0.91 | 0.79 | 1.06 |
|  | Saturday |  | – 0.05 | 0.07 | 0.95 | 0.83 | 1.10 |

\*: $p \leq 0.05$ ; \*\*: $p \leq 0.01$ ; \*\*\*: $p \leq 0.001$

Compared to the unadjusted odds presented in **Table 1**, the results in **Table 2** show the adjusted odds, given the relative effects of the other variables. At any point in **Table 2**, the odds can be considered while holding the other effects constant. For example, non-motorists are more than twice as likely to succumb in a crash compared to drivers. Additionally, non-motorists are about 80% more likely to die compared to all other outcomes within their own group. This calls for urgent and serious consideration into the safety of non-motorists, as has been shown elsewhere in literature (*5 – 9*).

The coefficients presented in **Table 2** reveal the impacts of each variable on injury severity. The coefficients at each cut point are estimated for the variable with relaxed proportionality constraints. The cut points are between mild and moderate, moderate and serious, serious and severe, and severe and maximal injury severities. Note that the coefficients are not valid across multiple levels and generalizations about the overall effect of the variable on injury severity cannot be made for variables with relaxed proportionality assumptions (*40, 41*). In other words, estimated coefficients are valid only at the cut points. Where the proportionality assumption holds, the same coefficient is estimated and valid across all cut points.

## 5. DISCUSSION

The model coefficients estimated and presented in **Table 2** show that the expectation of a more severe injury increases as the roadway class changes from Interstate to Local Roads. Patients injured on minor arterials, major collectors, and local roads are about 20% more likely to sustain a more severe injury. However, the odds ratio balloons to 1.6 on a minor collector. These findings place a higher emphasis on developing suitable countermeasures against crashes on road classes 4 through 7 with a promise of greater reduction in severe injuries and mortality.

Overall, passengers are slightly more likely to suffer severe injuries compared to drivers at each cut point level, however, the coefficients are not significant except for the serious injury level. On the other hand, non-motorists are significantly more likely to be more severely injured compared to drivers. The expectation of a more serious injury also scales more than proportionately, from about a third more likely to sustain moderate injuries compared to mild injuries to about 2.4 times as likely to suffer a fatality compared to severe injuries.

The results from the BAC category indicate that people impaired below the per se limit of 0.08 are about 45% more likely than unimpaired people from suffering a higher injury severity. However, people impaired above the per se limit are 27% more likely to suffer graver injuries compared to the previous group. Since the proportionality assumption for this variable was not relaxed, odds are cumulative over the lower order variable categories. It must be noted that these results do not indicate whether impairment lead to a crash or exacerbated the severity of a crash because not all impaired people were responsible for a crash, they may have been passengers in a vehicle.

All else remaining the same, females are about 25% less likely than males to suffer a more severe injury. However, increasing age predisposes people to suffer worse outcomes, with about 30% more chance compared to lower order severities. Moreover, older people are almost twice as likely to have a fatal outcome over severe injuries. Lastly, it is found that Sunday is the least safe day of the week, however, only Mondays and Tuesdays have a statistically significant contribution to lower injury severity. Other time variables like seasonality and time of day were examined but were found to be not significant at any level.

It is found that the partial proportional odds model presented in this study is better than the proportional odds and non-proportional odds models. The PPOM is less parsimonious than the POM, but more than the NPOM. As discussed, PPOM takes a parsimonious approach by permitting the independent variables that meet the proportional odds assumption to take the same coefficient for all injury severity levels, and other independent variables to vary between injury severity levels, thereby limiting loss of model accuracy and interpretability while retaining some immunity from overfitting. **Figure 4** shows a plot of the Akaike Information Criterion (AIC) and the Bayesian Information (Schwarz) Criterion (BIC) for the POM, PPOM, and NPOM. It can be seen that the PPOM model performs better than the other two models by AIC and about as well as the POM by BIC. It is to be noted that the POM requires fewer coefficients compared to the PPOM, and so should score lower BIC when all else is equal. Since the scores are almost equivalent, PPOM can be considered to represent the data better.

**FIGURE 4:**
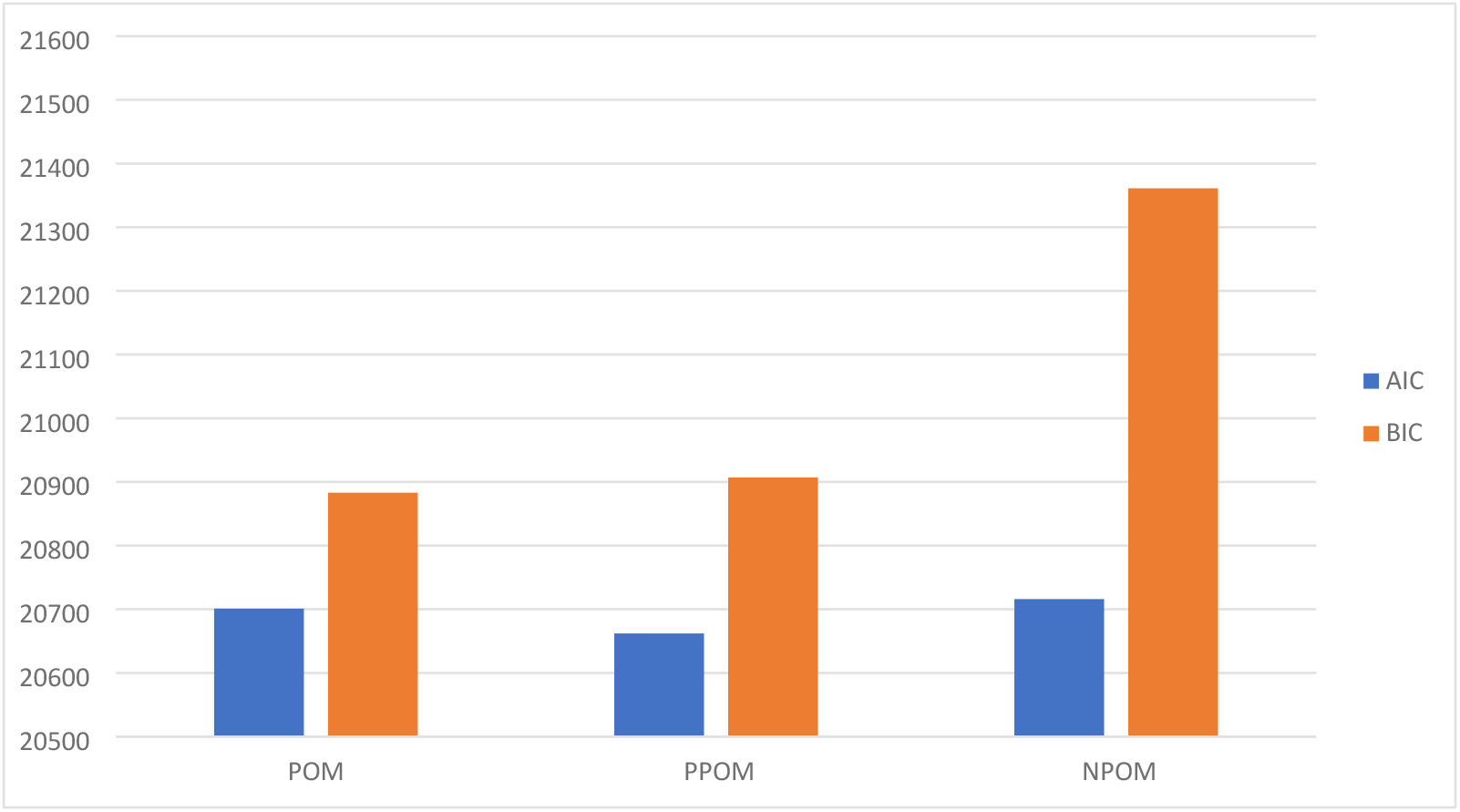
Akaike and Bayesian Information Criterion for POM, PPOM, and NPOM.

The results clearly show that as roadways become more local, injury severities and hospital outcomes worsen. This is a very worrying trend, especially considering that as roadways become more local, the number of vulnerable road users increases. On principal arterials, there could be the occasional unexpected pedestrian or bicyclist. However, on minor arterials, collectors, and local roads, the frequencies of pedestrians, bicyclists, micro-mobility vehicle users, older adults, and children are much higher. Further, rapid shifts in mode share, made possible by modern battery-operated micro-mobility vehicles, especially for the last mile, are only exacerbating the issue.

Majority of the current roads are ill-equipped to handle mixed use by pedestrians, bicyclists, electric micro-mobility vehicles, and traditional automobile traffic, often contributing to crashes where one party is unprotected outside a vehicle and can be seriously injured. Further, a growing number of older drivers are adding to the number of vulnerable road users as older people are more susceptible to serious injuries. Older drivers may also prefer traveling on local roads more than interstates (see interquartile ranges of age plot in **Figure 2**), be on medication that can impair judgement, have poorer reaction times due to age related deterioration of mental and physical capabilities, and other conditions, increasing the likelihood of being in or causing a crash with worser outcomes.

The data from the police and the hospital contain many other explanatory variables, such as safety equipment (seat belt, helmets, etc.) use, vehicle type, vehicle speeds, turning movements (including pedestrian movements), driver or non-motorist distractions, injuries in every body region, hospital lengths of stay, procedures performed, complications, preexisting conditions, and so on. Moreover, the researchers have access to a wealth of external datasets like admissions or visits to any hospital in Maryland, citations and adjudications, licensing and registration, weather, lighting conditions, pavement conditions, built environment and social activities, and so on. The presented model can be improved by including these data. However, these additional data sources tend to be noisier and more incomplete than those currently utilized in this paper, necessitating substantial data processing efforts prior to integration into comparable models.

## 6. CONCLUSION

The presented paper is the first work known to the authors that model hospital provided global consensus derived injury severity ranking with functional roadway classification and blood alcohol content obtained from a comprehensive blood test conducted at the hospital. Additionally, the study examines the effects of gender, age, race, day of week, and person type (driver, passenger, non-motorist) on injury severity. The results strongly suggest that infrastructure improvement emphasis should be placed on road classes from minor arterials to local roads to reduce severe and fatal injuries. Further, non-motorists are more than twice as likely to succumb in a crash compared to drivers and require urgent attention, especially given the recent increasing trend in their fatalities (*20, 21*).

This study is a preliminary look into how the built environment that favors automobile users is impacting the safety of all road users. The authors hope to examine the interplay between the built environment and conditions that negatively impact road user safety in future works. For example, the lack of suitable alternatives to driving may be the cause behind a lack of progress in reducing impaired driving crashes. Moreover, the impact of the rapidly changing technology available in vehicles contrasted with the increasing size of vehicles needs examination. The authors hope to build on the presented study to explore these and other interplays.

Lastly, this study has significant implications for the Safe System Approach (SSA). In its current form, the SSA calls for shared responsibility and improving the infrastructure, vehicles, individual behaviors, and response to crashes in an attempt to improve safety (*42, 43*). This study findings allow the application of efforts in a targeted manner by focusing on the most impactful aspects as determined by the presented coefficients.

## Data Availability

Data are available from the Injury Outcomes Data Evaluation System (IODES) at Charles “McC” Mathias National Study Center for Trauma and EMS (NSC) at the University of Maryland School of Medicine (UMSOM). Institutional Data Access / Ethics Committee (contact via IODES, Shock Trauma Center) for researchers who meet the criteria for access to confidential data.

## 7. DATA STATEMENT AND ACKNOWLEDGEMENT

This research was made possible by the Injury Outcomes Data Evaluation System (IODES) database maintained at the Charles “McC” Mathias National Study Center for Trauma and EMS (NSC) at the University of Maryland School of Medicine (UMSOM). IODES is a census of all injuries that occur in Maryland and maintains multiple different datasets that range to cover all aspects of injuries from the causing event, transport to the hospital, hospital care, and post-discharge. It also incorporates datasets beyond the injury, such as citations and adjudications, licensing and registration, roadway and built environment, and so on. The availability of these data in IODES allowed the analysis presented in this paper. Confidential data containing Personally Identifiable Information (PII) or Personal Health Information (PHI) in IODES are obtained under data use agreements with respective data owners. Confidential data include Shock Trauma Center registry data and the police crash data. Other datasets used are publicly available.

## 8. AUTHOR CONTRIBUTIONS

The authors confirm contribution to the paper as follows: study conception and design: K. Kaushik, A. Adegboye; data collection: K. Kaushik; analysis and interpretation of results: A. Adegboye, K. Kaushik; draft manuscript preparation: A. Adegboye, K. Kaushik. All authors reviewed the results and approved the final version of the manuscript. The authors further acknowledge the Practicum course at SOM that allowed the authors to develop the models presented in this paper. This work is unfunded.

---

Figure 3 shows plots of the empirical cumulative logit function as a visual guide of the parallel lines assumption.

